# Cost-effective serological test to determine exposure to SARS-CoV-2: ELISA based on the receptor-binding domain of the spike protein (Spike-RBD_N318-V510_) expressed in *Escherichia coli*

**DOI:** 10.1101/2020.09.15.20195503

**Authors:** Alan Roberto Márquez-Ipiña, Everardo González-González, Iram Pablo Rodríguez-Sánchez, Itzel Montserrat Lara-Mayorga, Luis Alberto Mejía-Manzano, José González-Valdez, Rocio Ortiz-Lopez, Augusto Rojas-Martinez, Grissel Trujillo-de Santiago, Mario Moisés Alvarez

## Abstract

Massive worldwide serological testing for SARS-CoV-2 is needed to determine the extent of virus exposure in a particular region, the ratio of symptomatic to asymptomatic infected persons, and the duration and extent of immunity after infection. To achieve this aim, the development and production of reliable and cost-effective SARS-CoV-2 antigens is critical.

Here, we report the bacterial production of the peptide S-RBD_N318-V510,_ which contains the receptor binding domain of the SARS-CoV-2 spike protein. We purified this peptide using a straightforward approach involving bacterial lysis, his-tag mediated affinity chromatography, and imidazole-assisted refolding. The antigen performances of S-RBD_N318-V510_ and a commercial full-length spike protein were compared in two distinct ELISAs. In direct ELISAs, where the antigen was directly bound to the ELISA surface, both antigens discriminated sera from non-exposed and exposed individuals. However, the discriminating resolution was better in ELISAs that used the full-spike antigen than the S-RBD_N318-V510._ Attachment of the antigens to the ELISA surface using a layer of anti-histidine antibodies gave equivalent resolution for both S-RBD_N318-V510_ and the full length spike protein.

Our results demonstrate that ELISA-functional SARS-CoV-2 antigens can be produced in bacterial cultures. S-RBD_N318-V510_ is amenable to massive production and may represent a cost-effective alternative to the use of structurally more complex antigens in serological COVID-19 testing.

## Introduction

The severe acute respiratory syndrome coronavirus 2 (SARS-CoV-2), the causal agent of the coronavirus disease 19 (COVID-19) has infected more than 28 million people^1^, at the time of this writing. Never before in contemporary history has humankind faced an infectious disease at this scale.

Massive worldwide serological testing is needed to determine the relevant epidemiological indicators related to COVID-19 infection, including the extent of the exposure, the ratio of symptomatic to asymptomatic infected persons, and the duration and extent of immunity after infection^2–5^. Moreover, as vaccines are developed, tested in animal models and humans, and applied to open populations, we will depend on assays for reliable and quantitative characterization of the immune responses associated with the administration of a vaccine to determine the level of immunization conferred^2,6^.

Fortunately, the time kinetics of the various antibodies (IgAs, IgMs, and IgGs) produced against SARS-CoV-2 in COVID-19 patients has been well described in recent reports^7^. For instance, we know that the determination of IgGs 15 days after viral exposure is a good indicator of a previous infection. Several semi-automated serological assays are commercially available to determine the likelihood of infection^8–10^. Most established commercial platforms perform well, in terms of accurate prediction of infection in convalescent patients, when the analysis is performed 15 days (or more) after a possible contact^10^. However, despite this reliability, automated serological platforms are expensive when compared to other techniques, such as regular enzyme-linked immunoassays (ELISAs).

ELISAs continue to be the most reliable and widely used method for characterization of the amount of antibodies developed against a specific antibody^10,11^. Laboratories around the world, and particularly in developing countries, depend on traditional ELISAS to conduct widespread serological testing. Therefore, reliable and cost-effective antigens for ELISA testing are greatly needed.

In the context of COVID-19 research, a limited number of reports have been published on the development and characterization of SARS-CoV-2 antigens for ELISAs ^12–15^. The Spike protein (S)^16,17^ and the nucleocapside protein (N)^18^ of SARS-CoV-2 have been used for COVID-19 serological diagnostic^13^. However, only a few detailed reports have been made available on the characterization of ELISAs for the identification of anti-SARS-CoV-2 antibodies.^5,12,13,19^ Most of these reports describe transient mammalian cell expression^12,14,20^ of the entire spike protein of SARS-CoV-2, or a fraction of the spike protein containing the RBD receptor binding domain^20^.

Here, we report the production of an antigen inspired by the structure of the receptor binding domain (RBD)^20^ of the spike protein of SARS-CoV-2. This antigen is produced by bacterial culture of *Escherichia coli*, which enables massive production at low cost^21^. In addition, we characterize and contrast the performance of two ELISA versions, involving (a) direct attachment of the antigen to the surface of plates or (b) the use of a bed of anti-histidine antibodies (anti-his–mediated ELISA) to engineer the reactive surface.

## Results and discussion

### Antigen design and production

We engineered an expression construct for the recombinant production of the RBD of the S protein of SARS-CoV-2. Specifically, we selected the region of the S-RBD between the residues N318 and V510 of the consensus sequence of the S protein of SARS-CoV-2.

In a recent report, a similar fraction of the SARS-CoV-2 spike protein (from residues 331 to 510) containing the RBD has been transiently expressed in HEK293 cells.^20^ That peptide successfully recognized the ACE receptor, the native target of the RBD of the spike protein^20^. Our construct also contained a region for the expression of M FH8, an enterokinase restriction site, as well as a histidine tag (his-tag). The M FH8 provides the means to facilitate purification and increase solubility of the product^22^.

The his-tag provides an additional handle for separation using his-tag affinity columns (loaded with divalent ions such as Ni^+2^). In addition, antigenic proteins containing histidine tags can be fixed to surfaces through anti-histidine antibodies to enable ELISA serological assays using his-tagged antigens^21,23^. Figure 1a schematically shows the sequence that we used to encode and produce the MFH8-RBD_Spike_-HisTag protein (S-RBD_N318-V510_ for short). This expression cassette was inserted in a plasmid for expression in *E. coli*. Figure 1c shows the molecular 3D structure of this product, as predicted by molecular structure simulations.

**Figure 1.**
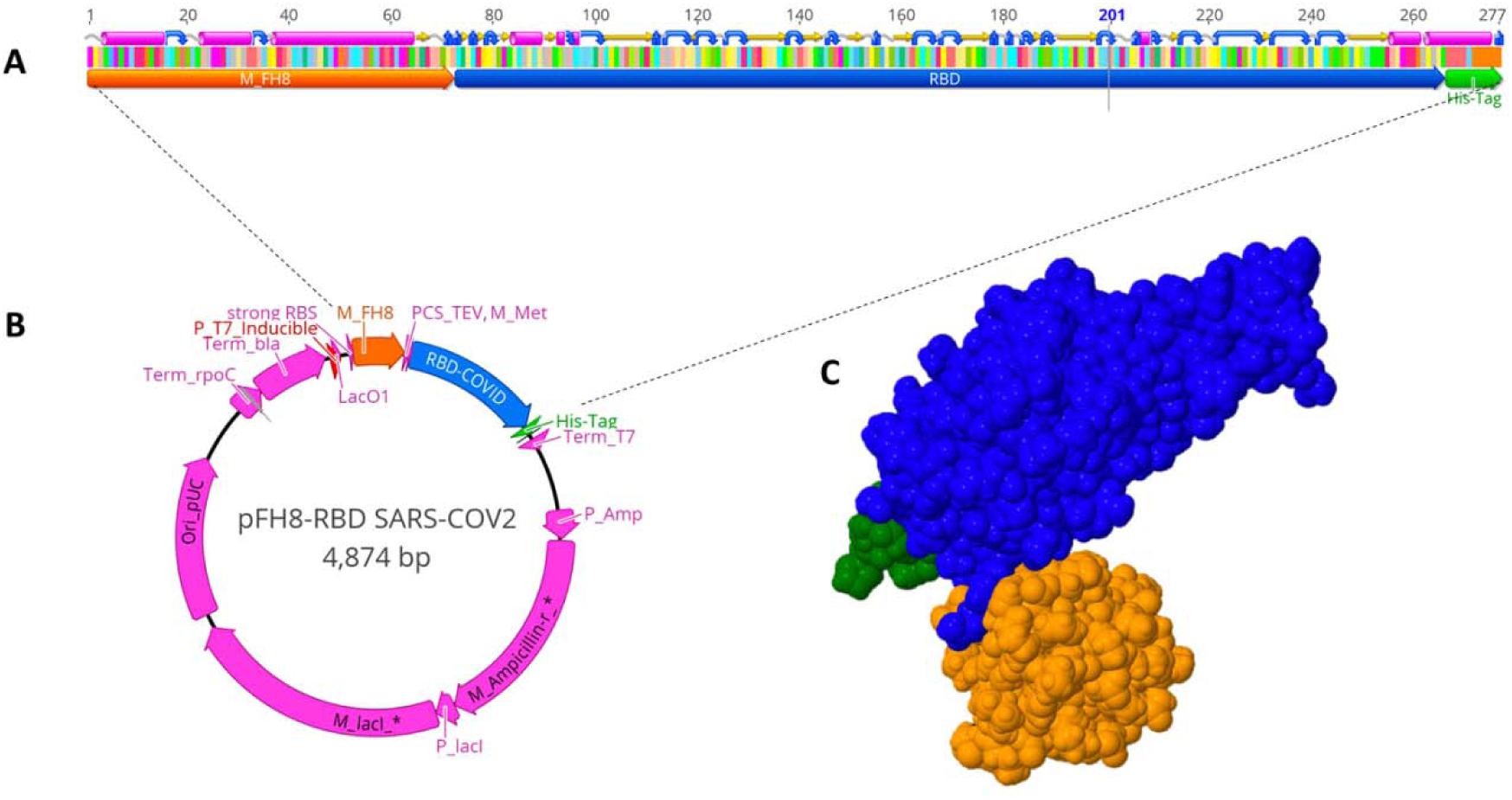
**Expression of** S-RBD_N318-V510_ in *Escherichia coli*. (A) Schematic representation of the sequence used to produce the MFH8-RBD_Spike_-HisTag protein (S-RBD_N318-V510_). This expression cassette was inserted into the (B) pFH-RBD SARS-CoV-2 plasmid for expression in *E. coli*. (C) Molecular 3D structure of the S-RBD_N318-V510_ protein, as predicted by molecular structure simulations.

We cloned the construct for the production of S-RBD_N318-V510_ in *E. coli* BL21 strain C41. High-producer clones were further cultured using Luria-Bertani (LB) medium in Erlenmeyer flasks and recombinant expression was induced using isopropyl β-d-1-thiogalactopyranoside (IPTG). Reproducible productions of up to 2 g (dry weight) of biomass L^-1^ was obtained in 2 L Erlenmeyer flasks incubated in orbital shakers at 30 °C for 12 h (Figure 2A). At this point, this production process has not yet been scaled up to an instrumented bioreactor. However, based on our previous work with other antigens expressed in bacterial systems, we anticipate that this scale-up will further increase biomass production to 10–20 g L^-1^.^24^

**Figure 2.**
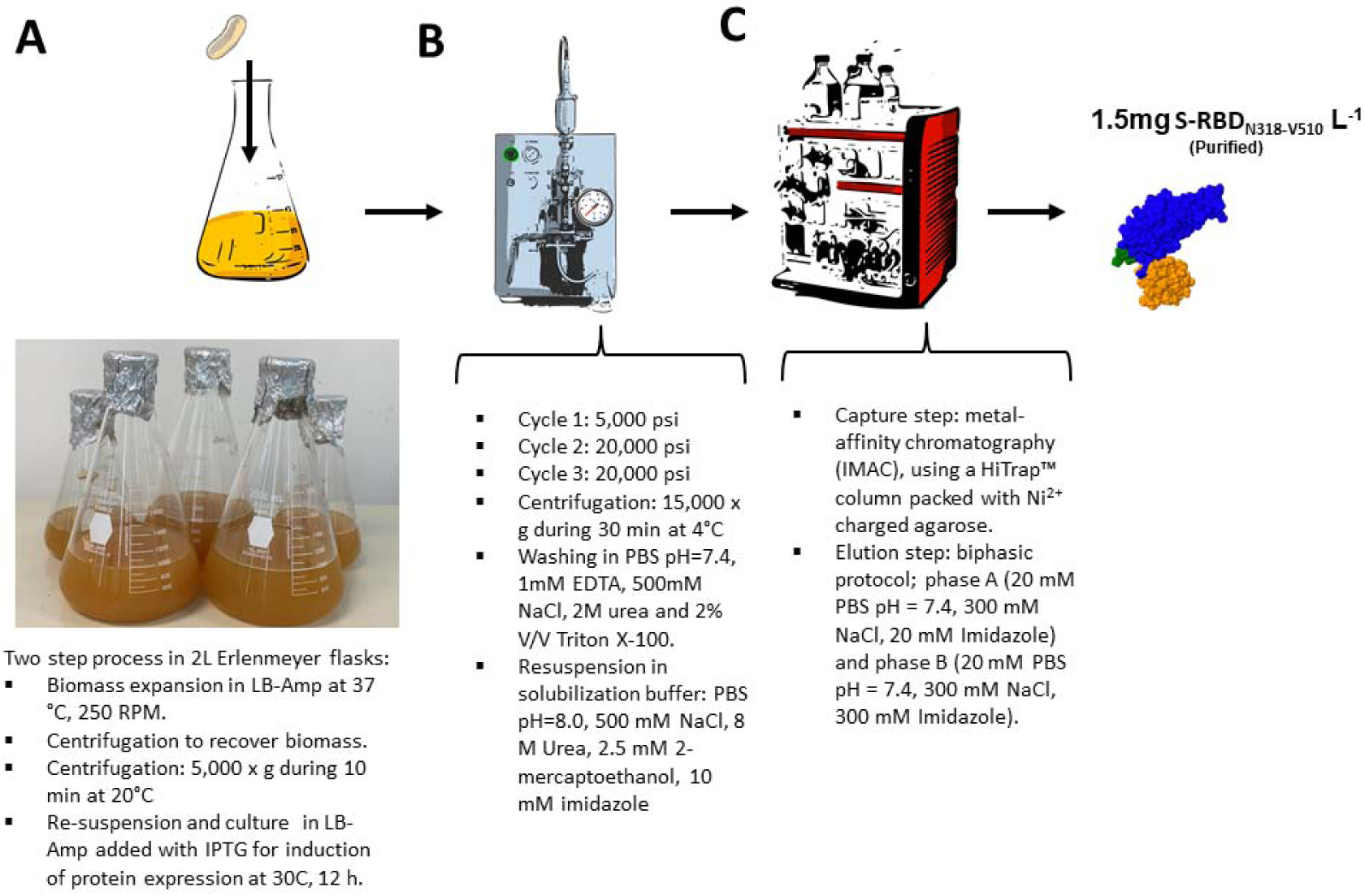
Production process for obtaining S-RBD_N318-V510_ from *Escherichia coli* cultures. Schematic representation of the general production process for **S-RBD**_**N318-V510**_. (A) Culture of a recombinant BL21 C41 strain of *E. coli* engineered to produce **S-RBD**_**N318-V510**_, (B) cell lysis in a continuous homogenizer, (C) and purification through several stages of affinity chromatography, in-column refolding, and elution.

### Recovery and purification

The methods described here lead to the production of S-RBD_N318-V510_ in inclusion bodies (IBs); we found negligible amounts of the protein in the supernatant of *E. coli* cultures. We implemented a conventional separation purification protocol that included lysis in a high-throughput homogenizer, filtration, re-suspension, and purification using his-tag columns (Figure 2). Lysis experiments were conducted in a high pressure homogenizer operated at 5000 Psi (first cycle) and 20,000 Psi in two subsequent cycles (Figure 2B).

After lysis, different refolding and purification strategies were tested, including the use of different columns and combinations of resuspension buffers and conditions. The best results were obtained by suspending the cell pellet in IB washing buffer at a ratio of 25 mL per g of IB pellet (wet weight), centrifuging to recover the pellet, washing with PBS, and re-suspending in IB solubilization imidazole-based buffer (Figure 2C).

The S-RBD_N318-V510_ protein was then purified by immobilization metal-affinity chromatography in a preparative chromatography system (Figure 2D). After testing different purification protocols, we opted for a two-phase purification protocol (as described in Materials and Methods).

Western blots conducted using marked anti-histidine antibodies indicated that the recombinant product was produced and could be purified with sufficient yield and purity by the methods described here. The molecular weight of the S-RBD_N318-V510_ protein (approximately 31 kDa) was consistent with the expected value. The degree of purity of the RBD was estimated at approximately 92% based on the SDS-PAGE protein profiles and using the Image J open source software for scanning densitometry analysis. We have consistently obtained overall yields of approximately 1.5 mg of pure S-RBD_N318-V510_ per liter of culture medium among different batches.

### Determination of binding affinity

We evaluated the binding affinity of the S-RBD_N318-V510_ protein in two sets of ELISA experiments using commercial anti-RBD antibodies.

In the first set of experiments (direct ELISAS; Figure 3A), we directly deposited 1 µg of purified S-RBD_N318-V510_ per well in 96-well plates. We then added a commercial anti-S(RBD) antibody to each well, and the relative amount of antibody bound to S-RBD_N318-V510_ protein was determined by absorbance after the addition of an anti-heavy chain antibody marked with horseradish peroxidase (HRP). The S-RBD_N318-V510_ protein exhibited a binding affinity of approximately 75.51 ± 5% of that of the commercial control (Figure 3B).

**Figure 3.**
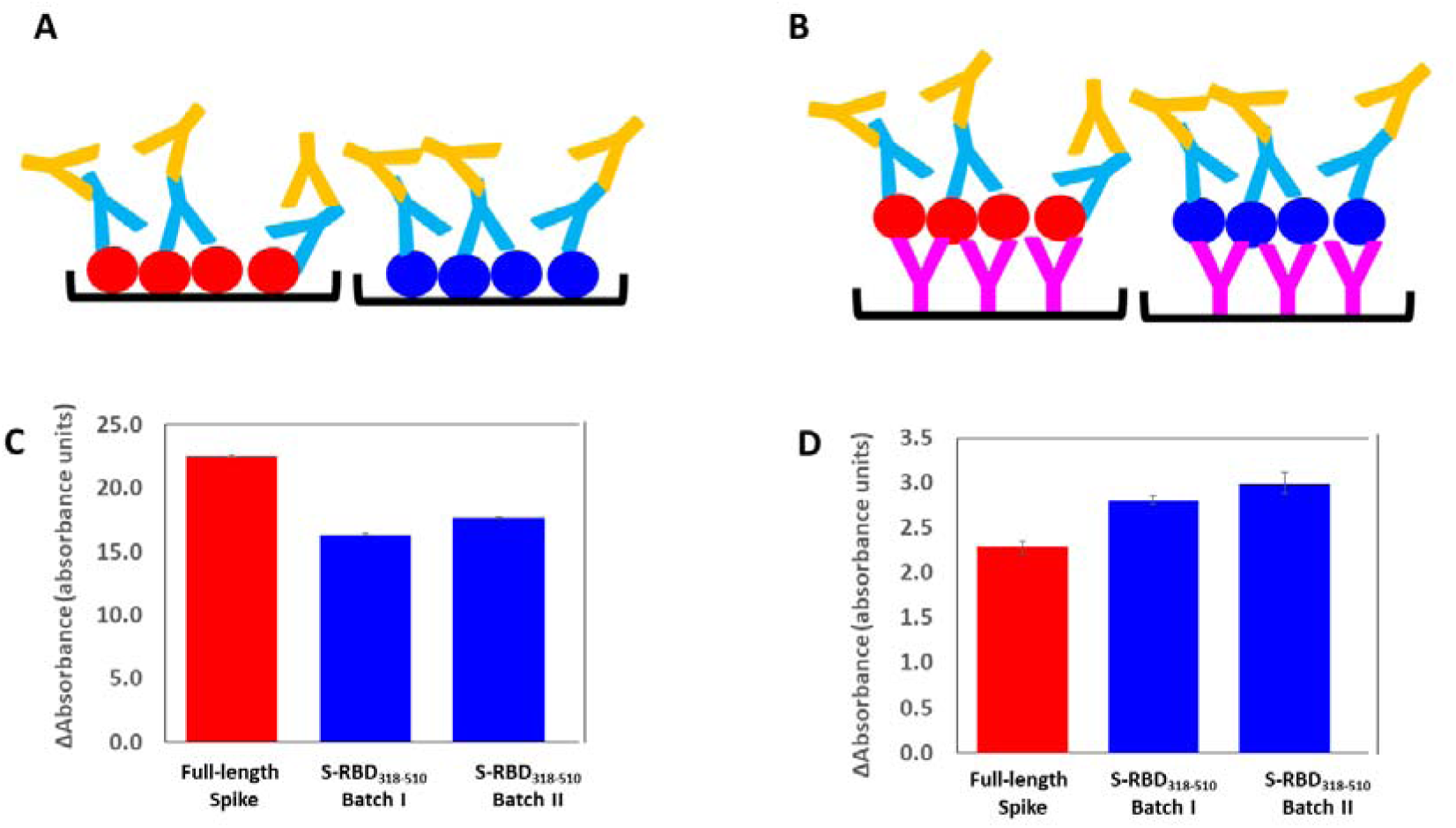
Contrast of two versions of ELISAs for identification of anti-Spike SARS-CoV-2 antibodies. Comparison of a commercial full-length spike protein from SARS-CoV-2 (red circles) and the S-RBD_N318-V510_ protein (blue circles) produced in *Escherichia coli* as antigens in ELISA experiments performed to identify anti-spike SARS-CoV-2 antibodies. The antigen was (A) directly bound to the surface of 96-well plates, or (B) bound through a layer of anti-histidine antibodies (pink Ys). In both cases, the specific attachment of anti-spike SARS-CoV-2 antibodies (blue Ys) was revealed by binding of anti-human heavy chain antibodies functionalized with horse-radish peroxidase (yellow Ys). Comparison of absorbance readings for (C) direct ELISAs or (D) anti-histidine mediated ELISAs. A commercial full-length spike (red bars) or the S-RBD_N318-V510_ protein (blue bars) were used as antigens. Commercially available anti-spike SARS-CoV-2 antibodies were used as reactants.

In the second set of experiments, we conducted sandwich-type ELISAs (Figure 3C). For that purpose, the S-RBD_N318-V510_ protein was bound to the bottom surface of the 96-well plates through an anti-histidine antibody.^21^ In concept, this strategy may promote a more uniform orientation of the S-RBD_N318-V510_ protein and thereby improve the selectivity of the assay. Indeed, the binding ability of S-RBD_N318-V510_ protein was 26.63 ± 5% higher than that shown by the commercially available spike protein used here as a positive control (Figure 3D).

### Determination of binding affinity using human sera

We ran an additional series of ELISA experiments using actual human sera and contrasted the results with those of the two ELISA versions previously discussed.

In a first round of experiments, we directly bound commercial spike protein or S-RBD_N318-V510_ protein to 96-well ELISA plates. First, we used 5 serum samples from non-exposed individuals collected from June to December 2009 during the first wave of pandemic Influenza A/H1N1/2009 in México. The average absorbance value exhibited by samples of these non-exposed individuals was 0.272 (99% CI 0.243 to 0.301) in ELISAs conducted using the S-RBD_N318-V510_ protein. Similarly, the average absorbance value for non-exposed individuals when the whole spike protein was used was 0.198 (99% CI 0.168 to 0.224). Sera from non-exposed individuals exhibited low absorbance values and enabled the definition of an average reliable absorbance value for non-exposed individuals (first two bars in Figure 4A,B). Figure 4A and 4B show the absorbance readings from direct ELISA experiments conducted on a set of selected serum samples. In this set, we included sera from non-exposed COVID-19 individuals (the samples collected in 2009). We also selected samples from probably exposed individuals that exhibited absorbance values statistically similar to negative samples, as well as sera from convalescent patients diagnosed as COVID-19 (+) by RT-qPCR. The performance of the S-RBD_N318-V510_ protein (blue bars) and a commercial spike protein (magenta bars) was compared. Some of the samples exhibited values that exceeded the thresholds of the 99.5% confidence values for serologically negative samples. Consistently, these samples corresponded to sera from convalescent COVID-19 patients.

**Figure 4.**
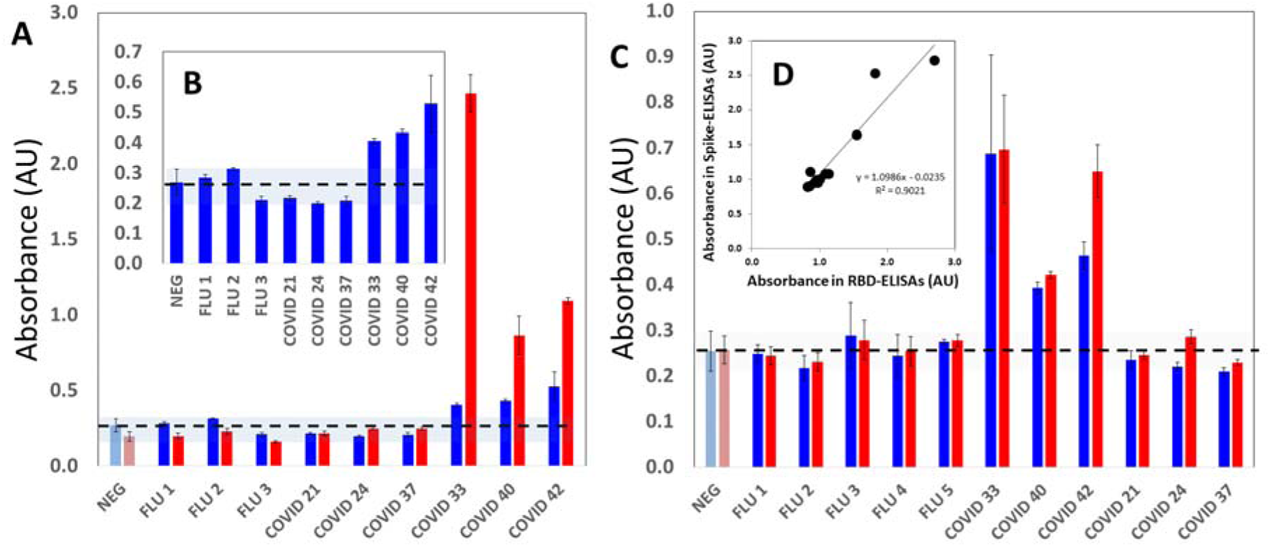
Binding of antibodies from human sera to S-RBD_N318-V510_ using direct and anti-histidine–mediated ELISAs. (A) Binding of antibodies from human sera, measured as absorbance readings, in direct ELISA experiments that used a commercial full-length spike (red bars) or the S-RBD_N318-V510_ protein (blue bars) as antigens. Serum samples were obtained from non-exposed volunteers (FLU X; collected during pandemic Influenza A/H1N1/2009) and from volunteers possibly exposed to SARS-CoV-2 (COVID X; collected during pandemic COVID-19). NEG bars indicate the average absorbance reading exhibited by serum samples from non-exposed volunteers. (B) Absorbance readings in direct ELISA experiments that used the S-RBD_N318-V510_ protein as the antigen. (C) Binding of antibodies from human sera, measured as absorbance readings, in anti-histidine mediated ELISA experiments that used a commercial full-length spike (red bars) or the S-RBD_N318-V510_ protein (blue bars) as antigens. Serum samples were obtained from non-exposed volunteers (FLU X; collected during pandemic Influenza A/H1N1/2009) and from volunteers possibly exposed to SARS-CoV-2 (COVID X; collected during pandemic COVID-19). NEG bars indicate the average absorbance reading exhibited by serum samples from non-exposed volunteers. (D) Graphic analysis of the correlation between titers obtained in anti-histidine– mediated ELISAs that used the commercial full-length spike (red bars) or the S-RBD_N318-V510_ protein (blue bars) as antigens. All experiments were conducted using 1:100 serum dilutions.

Both the full-length commercial spike antigen and the S-RBD_N318-V510_ protein antigen were able to discriminate between samples from non-exposed individuals and COVID-19 (+) patients. However, consistent with recent reports^25^, the absorbance values were much higher when the full-length commercial spike protein was used as the ELISA antigen than when the S-RBD_N318-V510_ protein was used. Therefore, in this ELISA format, the difference in absorbance value between positive and negative samples was greater when the spike protein was used than when the S-RBD_N318-V510_ protein is used.

We repeated these ELISA experiments using a second strategy consisting of binding the antigens through a layer of anti-histidine antibodies. Figure 4C shows the results of a parallel ELISA experiment using a layer of anti-histidine antibodies to bind the antigens to the plate surfaces.

The average absorbance value for non-exposed individuals was 0.257 (99% CI 0.237 to 0.277) and 0.255 (99% CI 0.226 to 0.284) for the full-length spike and the S-RBD_N318-V510_ protein, respectively. In general, the absolute values of the absorbance readings were lower in the anti-histidine–mediated ELISAs than in the direct ELISAs. The absorbance readings produced using the full spike and S-RBD_N318-V510_ were remarkably similar (Figure 4C and D). This suggests that the anti-histidine antibodies allow similar arrangement and alignment of both antigens to present reactive surfaces of comparable antibody binding capacity.

We further studied the usefulness of these two ELISA versions (direct or mediated by anti-histidine antibodies) by testing 50 samples from convalescent patients diagnosed as COVID-19 (+) by RT-qPCR. Most of these patients had shown COVID-19–related symptoms at least 7 days before blood collection. Figure 5A shows the normalized absorbance readings for this set of serum samples, with no particular order, as determined by ELISA testing conducted by direct sensitization of the reactive surface with S-RBD_N318-V510_. Figure 5B shows results from ELISAs that used an anti-histidine–mediated binding to sensitize the reaction with S-RBD_N318-V510_. The normalization of the absorbance values consisted of dividing the absolute absorbance by the average value of absorbance readings of sera from three non-exposed individuals.

**Figure 5.**
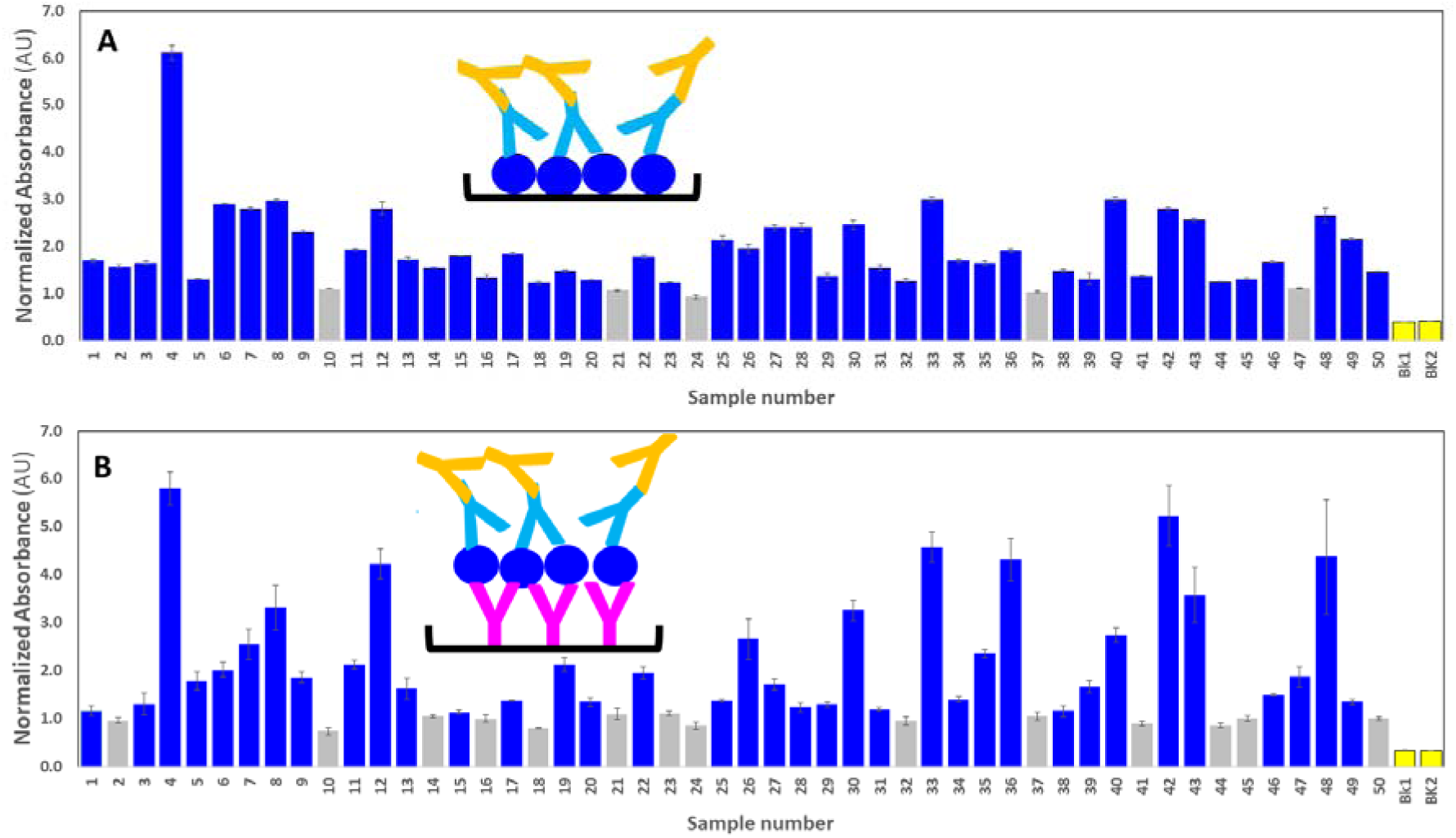
Binding of S-RBD_N318-V510_ serum samples donated by convalescent patients confirmed as COVID-19 (+) by RT-qPCR. Normalized absorbance readings related to the binding affinity of the S-RBD_N318-V510_ protein to human sera antibodies from COVID-19 convalescent patients for (A) direct, and (B) anti-histidine mediated ELISAs. All absorbance readings were normalized by the average absorbance reading exhibited by samples from non-exposed individuals. Normalized readings higher than the threshold value of 1.11 (for direct ELISAs) or 1.12 (for anti-histidine–mediated ELISAs) are indicated in blue. Absorbance readings from blanks (phosphate buffered saline only) are indicated in yellow as a reference. Experiments were conducted using 1:100 serum dilutions.

We also established threshold values for normalized absorbance to discriminate between the negative and positive results for threshold values of normalized absorbance of 1.11 and 1.12 for direct and anti-histidine mediated ELISAS using S-RBD_N318-V510_, respectively. These values were slightly above the upper threshold of the 99% CI for readings of non-exposed individuals.

The results of both ELISA formats were highly consistent (Figure %A-C). As shown earlier, anti-his–mediated ELISAs yielded similar results, regardless of the use of the full-length spike protein or S-RBD_N318-V510._ Therefore, we assumed that the anti-his–mediated results correlated well with ELISAs conducted with the full spike protein and can be taken as a reference for determining the sensibility and specificity of direct ELISAs performed using S-RBD_N318-V510._ When this is done, the selectivity and specificity of the direct S-RBD_N318-V510_ format are 97.2% and 52.0 %, respectively, when a threshold value of normalized absorbance of 1.10 is used. If a threshold value of normalized absorbance of 1.25 is used instead, the values of selectivity and specificity are 97.2% and 68.0%, respectively. The overall accuracy of the direct S-RBD_N318-V510_ ELISA test (i.e., the overall consistency of the results with respect to the anti-his S-RBD_N318-V510_ ELISA) was 81.8% and 85.45% when the thresholds were set at 1.10 and 1.25, respectively.

Overall, the results suggest that the anti-his S-RBD_N318-V510_ ELISA is more consistent with full-length spike ELISAS. However, direct S-RBD_N318-V510_ ELISA can be used in serological testing (further reducing the cost) with only a minimum sacrifice of selectivity, but with an increased probability of false positives.

## Conclusions

Arguably, antigens are one of the most important reagents that clinicians will require while facing COVID-19 pandemics in the months to come. Here, we report methods for the production of a portion of the S1 fraction of the SARS-CoV-2 spike protein that contains the receptor-binding domain for the Angiotensin II human receptor.

We chose *Escherichia coli* as an expression host, and we describe a straightforward process, amenable to widespread implementation, for the production and purification of the S-RBD_N318-V510_ protein. Our aim was to enable the widespread use of this simple process to produce a cost-effective SARS-CoV-2 antigen. In ELISA experiments using commercial anti-spike antibodies or actual sera from patients, this protein performs similarly to commercially available antigens based on the expression of larger segments of the spike protein.

The fact that our antigen is expressed in bacterial systems greatly facilitates its production and paves the way to scaling up. We show that antigen production of 1.5 mg per L is feasible, even when using non-agitated Erlenmeyer flasks and non-instrumented bioreactors. This production level is already attractive, since production can be completed in 24 h. Complete COVID-19 ELISA kits are commercially available but their cost (approximately 8 USD per reaction per well) still limits massive implementation, particularly in developing economies. The current value of commercially available S1-derived SARS-CoV-2 antigens is approximately 7 USD µg^-1^, which is also prohibitive for most laboratories for large-scale screening of COVID-19 seropositive subjects. We believe that a lab-scale manufacturing operation based on the process described here may allow the production of gram amounts of antigen per month of satisfactory quality to enable mass-scale screening projects in open populations.

## Materials and methods

### Design of S-RBD_N318-V510_ and prediction of its 3D structure

We used the Geneious 11.1.5 software (Biomatters, Ltd., New Zealand) to design the vector pFH8-RBD SARS-COV2 that contained the RBD (region of 193 aa from N318-V510) of the SARS-CoV-2 spike protein. Histidine and FH8 tags were added for use in the purification process. The 3D structure of the RBD protein with tags was predicted using the software I-TASSER server (University of Michigan, USA).

### Cloning and transformation

The full spike coding sequence was synthesized by Genscript (New Jersey, USA) and was used to obtain the sequences comprising the RBD. This sequence was cloned in an expression vector (ATUM, CA, USA) regulated by a T7 promoter (IPTG-inducible) using a SapI restriction site and T4 DNA ligase (New England Biolabs, UK). The expression vector was transformed into chemical-competent *E. coli* C41 cells (Lucigen Corporation, WI, USA) to obtain a producer clone.

### RBD production in Erlenmeyer flasks

The highest producer clone was cultured in Luria-Bertani broth containing 50 µg/mL ampicillin (LB-Amp) in 2L Erlenmeyer flasks. For the initial growth, 200 mL of LB-Amp broth was maintained overnight at 37°C with 250 rpm agitation in an orbital shaker (VWR International, USA). After 12 h of culture, cells were harvested using a Z36 HK centrifuge (Hermle Labortechnik, Germany) at 5000 × g for 10 min. The cell pellet was then resuspended in fresh LB-Amp broth containing 1 mM isopropyl β-d-1-thiogalactopyranoside (IPTG) to induce RBD production. The induction was conducted at 30°C with agitation at 250 rpm for 8–12 h. After induction, cells were recovered by centrifugation at 5000 × g for 10 min at 4°C. Cell pellets were kept at -20°C until further processing.

### S-RBD_N318-V510_ recovery and purification

Pellets from IPTG-induced cells were re-suspended in PBS buffer (pH=7.4) containing 100 mM NaCl in a proportion of 7.5 mL per gram of cells (wet weight). The cells were then disrupted in an EmulsiFlex-C3 high-pressure homogenizer (Avestin, Canada). The process comprised 3 cycles, with the first cycle set to reach 5000 psi and the following two cycles performed at 20000 psi. Cell lysates were centrifuged at 15,000 × g for 30 min at 4°C in a Z36 HK centrifuge. The pellet containing the IB was re-suspended in IB wash buffer (PBS, pH=7.4, 1mM EDTA, 500 mM NaCl, 2 M urea, and 2% Triton X-100) at a ratio of 25 mL per g of IB pellet (wet weight), centrifuging to recover the pellet, washing with PBS, and re-suspending in IB solubilization buffer (PBS, pH=8.0, 500 mM NaCl, 8 M Urea, 2.5 mM 2-mercaptoethanol, and 10 mM imidazole) (Figure 2C).

The S-RBD_N318-V510_ protein was then purified by immobilization metal-affinity chromatography in a preparative chromatography system (Figure 2D). After testing different purification protocols, we opted for a two-phase purification protocol. Phase A consisted of 20 mM PBS, 300 mM NaCl, and 20 mM imidazole, pH=7.4, and phase B was 20 mM PBS pH = 7.4, 300 mM NaCl, and 300 mM imidazole at pH=7.4. The purification protocol was set with an initial equilibrium of 10 column volumes (CV) of phase A and a flow rate of 1 mL/min. A 5 mL sample of protein was injected at a flow rate of 0.5 mL/min. After sample injection, a washing step of 8 CV was set at a flow rate of 1 mL/min, followed by elution with 3 CV of a linear gradient from 0 to 80% of phase B, then 10 CV of 20/80% phase A/B, and finally 5 CV of 100% phase B, all at a flow rate of 1 mL/min. Finally, to prepare for further purifications, the column was re-equilibrated with 5 CV of phase A at 1 mL/min. The fraction containing the protein of interest was recovered based on the chromatogram and stored at 4°C.

This suspension was vigorously washed for 30 min at room temperature and then centrifuged at 15,000 × g for 30 min. The resultant pellet was gently washed with PBS to eliminate the excess Triton X-100 and then resuspended in IB solubilization buffer. This suspension was then vigorously stirred overnight at room temperature and finally centrifuged at 15,000 × g for 30 min at 4°C. The supernatant containing the solubilized IBs was recovered, filtered through a 0.2 µm syringe filter, and stored at 4°C.

The S-RBD_N318-V510_ protein was purified by immobilized metal-affinity chromatography (IMAC), using a HiTrap™ column (GE Healthcare, UK) packed with 1 mL Ni^2+^ charged agarose Ni-NTA Superflow (Quiagen, Germany) in an ALJkta Pure system (GE Healthcare, UK) chromatography system,. The degree of purity of S-RBD_N318-V510_ was estimated from SDS-PAGE protein profiles using Image J, an open source software for scanning densitometry analysis.

A dual phase separation strategy was implemented. Phase A consisted of 20 mM PBS, 300 mM NaCl, and 20 mM imidazole, pH=7.4, and phase B was 20 mM PBS pH = 7.4, 300 mM NaCl, and 300 mM imidazole at pH=7.4. The purification protocol was set with an initial equilibrium of 10 column volumes (CV) of phase A and a flow rate of 1 mL/min. A 5 mL sample of protein was injected at a flow rate of 0.5 mL/min. After sample injection, a washing step of 8 CV was set at a flow rate of 1 mL/min, followed by elution with 3 CV of a linear gradient from 0 to 80% of phase B, then 10 CV of 20/80% phase A/B, and finally 5 CV of 100% phase B, all at a flow rate of 1 mL/min. Finally, to prepare for further purifications, the column was re-equilibrated with 5 CV of phase A at 1 mL/min. The fraction containing the protein of interest was recovered based on the chromatogram and stored at 4°C.

### ELISA assays

We developed and characterized two ELISA strategies for the evaluation of presence of specific anti-SARS-CoV-2 antibodies (Figure 3), as described in the Results and Discussion. Standard commercial 96-wells micro-assay plates (CorningH, Maxisorp™; USA) were used.

In the first format, 100 µL of 1 µg/mL RBD in PBS was dispensed into each well and incubated for 8 h at 4°C, followed by addition of 100 µL 5% skim milk in PBS and further incubation for 1 h at room temperature, and then three washes with PBS containing 0.05% Tween-20™. Rabbit anti-COVID-19 pAb (100 µL; 1:2000 dilution; Sino Biological Inc., PA, USA) was then added incubated for 1 h at room temperature, followed by three washes with PBS containing 0.05% Tween-20™. The presence of rabbit antibodies was revealed by adding donkey anti-rabbit-HRP (100 µL, 1:5000 dilution; Pierce, Rockford IL, USA), followed by three washes with PBS containing 0.05% Tween-20™. The HRP was then detected by adding 100 µL 1-Step™ Ultra TMB-ELISA (Pierce, Rockford IL, USA) until a blue color was observed. The reaction was stopped by adding 100 µL 1M H_2_SO_4_ and the absorbance was measured at 450 nm in a Biotek microplate reader (VT US).

The second ELISA format consisted of first sensitizing the plate wells with mouse anti-histidine pAb (100 µL, 1:1000 dilution; Bio-Rad Laboratories, Inc., CA, USA) and incubating for 8h at 4°C, then blocking with skim milk for 1 h at room temperature, followed by 3 washes with PBS containing 0.05% Tween-20™. The plates were then incubated with RBD for 1 h at room temperature. All subsequent steps were as described for the first ELISA format.

### ELISA testing of serum samples

We performed ELISA experiments using samples of sera from non-exposed individuals and convalescent positive volunteers. Five samples of sera from COVID-19 non-exposed individuals were collected from volunteers at Hospital San José (Nuevo León, México), from June 2009 to October 2009, during pandemic Influenza A/H1N1/2009. Fifty-five samples of sera from convalescent patients previously confirmed as COVID-19 (+) by RT-qPCR were collected at Alfa Medical S.A. de C.V. (Monterrey, N.L., México). Samples were collected from patients after obtaining informed and signed written consent and in complete observance of good clinical practices, the principles stated in the Helsinki Declaration, and applicable lab operating procedures at Hospital Alfa. Every precaution was taken to protect the privacy of sample donors and the confidentiality of their personal information. The experimental protocol was approved on May 20th, 2020 by a named institutional committee (Alfa Medical Center, Research Comitte; resolution AMCCI-TECCOVID-001).

As described in the Results and Discussion section, the two different ELISA strategies were contrasted. In the first format, 100 µL of 1 µg/mL RBD in PBS was dispensed into each well of 96-well plates and incubated for 8 h at 4°C, followed by blocking with 100 µL 5% skim milk in PBS and incubation for 1 h at room temperature, and 3 washes with PBS containing 0.05% Tween-20™. Different dilutions (1:5, 1:50, 1:100 and 1:200; 100 µL) of serum from volunteers were added per well, incubated for 1 h at room temperature and then washed three times with PBS containing 0.05% Tween-20™. Best results were observed when 1:100 dilutions were used. The presence of human IgG was detected by adding goat anti-human IgG HRP (100 µL; 1:10000 dilution; Pierce Biotechnology Inc., IL, USA) and incubating for 1 h at room temperature, followed by three washes with PBS containing 0.05% Tween-20™ and detection with 1-Step™ Ultra TMB-ELISA

In the second format, RBD (100 µL; 1 µg/mL in PBS) was dispensed in each well and incubated for 1 h at room temperature followed by 3 washes with PBS containing 0.05% Tween-20™ .Then 100 µL of different dilutions (1:5, 1:50, 1:100, and 1:200) of serum from volunteers were added per well, incubated for 1 h at room temperature and subsequently washed three times with PBS containing 0.05% Tween-20™The presence of human IgG was again detected with goat anti-human IgG HRP and the remaining steps were conducted as described for the first ELISA format.

## Data Availability

All data related to this manuscript is contain within.

## Acknowledgements

The authors aknowledge the funding provided by the Federico Baur Endowed Chair in Nanotechnology (0020240I03). EGG acknowledges funding from a doctoral scholarship provided by CONACyT (Consejo Nacional de Ciencia y Tecnología, México). GTdS and MMA acknowledge the institutional funding received from Tecnológico de Monterrey (Grant 002EICIS01). MMA, GTdS, and IMLM acknowledge funding provided by CONACyT (Consejo Nacional de Ciencia y Tecnología, México) through grants SNI 26048, SNI 256730, and SNI 1056909, respectively.

